# Genetic variations in *EIF2AK3* are associated with neurocognitive impairment in people living with HIV

**DOI:** 10.1101/2022.04.20.22273997

**Authors:** Cagla Akay-Espinoza, Sarah Bond, Beth A. Dombroski, Asha Kallianpur, Ajay Bharti, Donald R. Franklin, Gerard D. Schellenberg, Robert K. Heaton, Igor Grant, Ronald J. Ellis, Scott L. Letendre, Kelly L. Jordan-Sciutto

## Abstract

Coding and noncoding single-nucleotide variants (SNVs) of *EIF2AK3*, which encodes an integrated stress response (ISR) kinase, may play a role in neurodegenerative disorders. We used a candidate gene approach to determine the correlation of *EIF2AK3* SNVs with neurocognitive (NC) impairment (NCI), which can persist with viral suppression from antiretroviral therapy (ART) in people with HIV (PWH). This retrospective study of prospectively collected data included participants of the CNS HIV Anti-Retroviral Therapy Effects Research (CHARTER) cohort, after excluding participants with severe neuropsychiatric comorbidities. Genome-wide data previously obtained in the CHARTER cohort participants (n=1,047) were analyzed to interrogate the association of three noncoding *EIF2AK3* SNVs with the continuous global deficit score (GDS) and global NCI (GDS≥0.5). Targeted sequencing (TS) was performed in 992 participants with available genomic DNA to determine the association of three coding *EIF2AK3* SNVs with GDS and NCI. Analyses included univariable and multivariable methods such as analysis of variance and regression. Multivariable models covaried demographic, disease-associated, and treatment characteristics. The cohort characteristics were as follows: median age, 43.1 years; females, 22.8%; European ancestry, 41%; median CD4+ T cell counts, 175/µL (nadir) and 428/µL (current). At first assessment, 70.5% used ART and 68.3% of these had plasma HIV RNA ≤ 200 copies/mL. A minority of participants had at least one risk allele for rs6739095 (T,41.7%), rs1913671 (C,41.4%), and rs11684404 (C,39.4%). All three noncoding *EIF2AK3* SNVs were associated with significantly worse GDS and more NCI (all *p*<0.05). By TS, fewer participants had at least one risk allele for rs1805165 (G,30.9%), rs867529 (G,30.9%), and rs13045 (A,41.2%). Homozygosity for all three coding SNVs was associated with significantly worse GDS and more NCI (all *p*<0.001). By multivariable analysis, the rs13045 A risk allele, current ART use, and Beck Depression Inventory-II (BDI) > 13 were independently associated with GDS and NCI (*p*<0.001). The other two coding SNVs did not significantly correlate with GDS or NCI after including rs13045 in the model. The coding *EIF2AK3* SNVs were specifically associated with worse performance in executive functioning, motor functioning, learning, and verbal fluency. Coding and non-coding SNVs of *EIF2AK3* were associated with global NC and domain-specific performance. The effects were small-to-medium in size but were present in multivariable analyses. Specific SNVs in *EIF2AK3* may be an important component of genetic vulnerability to NC complications in PWH. Identification of host factors that predict NCI could allow for earlier interventions, including those directly modulating the ISR, to improve NC outcomes.

## Introduction

Despite vastly improved health outcomes with antiretroviral therapy (ART), up to 50% of people with human immunodeficiency virus (HIV) infection (PWH) continue to experience neurocognitive (NC) impairment (NCI) and other neurological disorders ^1; 2^. Milder forms of NCI persist and may worsen even when HIV RNA is suppressed by ART ^2-5^. Progression to more disabling NC disorders is more likely in PWH with mild, even asymptomatic, NCI than in unimpaired PWH ^6^. Accumulating evidence also supports that neuropsychiatric disorders such as depression and aging-related disorders are more common in PWH ^7-14^.

Suppressive ART is currently the only proven intervention to prevent or improve NCI in PWH ^15-17^. Several factors increase the risk for NCI, including lower nadir CD4^+^ T cell count, metabolic syndrome, hepatitis C virus (HCV) coinfection, depression, and older age ^17-28^. To date, clinical trials have not identified interventions that further improve on the benefits of ART. Mechanistic biomarkers of NC vulnerability should enable earlier diagnosis of NCI and new therapeutic interventions ^29^.

While some of the risk factors associated with NCI, such as sex and genetic factors, may be known at the time of diagnosis and can be considered time-invariable, others, such as metabolic syndrome, arise later and can vary over time. While knowledge of risk factors is paramount for the successful prevention and management of NCI, identification of host factors that are predictive for these at the time of, or soon after, HIV diagnosis, could have an impact by providing opportunities to implement adjunctive therapies. Conversely, genetic vulnerability is well studied in several neurodegenerative disorders, but less is known about the potential contribution of genetic risk factors to HIV-associated NCI ^30-37^.

Several of the risk factors for NCI, including HIV replication, older age, and persistent inflammation, induce the integrated stress response (ISR), a ubiquitous cellular response pathway used by cells in the brain and elsewhere to resolve cellular stress and reach homeostasis ^38-51^. The ISR involves the activation of at least one of four sensor kinases, GCN2, HRI, PKR, and PKR-like endoplasmic reticulum kinase (PERK), in response to amino acid or heme deprivation, viral infection, and endoplasmic reticulum (ER) stress, with subsequent activation via multimerization and autophosphorylation. When active, the common target of all four ISR kinases is eukaryotic initiation factor 2α (eIF2α) which, when phosphorylated, attenuates global protein synthesis while upregulating the translation of a subset of transcripts involved in establishing protein homeostasis, such as ATF4 and genes involved in the antioxidant response ^39; 52; 53^. When ISR activation is prolonged, genes involved in cell death including caspase 4 and CCAAT/enhancer-binding proteins (C/EBP) homologous protein (CHOP) can be induced ^38; 39; 54-56^; thus, transient ISR activation may be protective but chronic activation may result in cell damage or death ^57-61^. Importantly, we and others have shown ISR activation, including the increased expression of phospho-PERK and phospho-eIF2 α (peIF2α), to be present in PWH with NCI as well as in people without HIV who have Alzheimer’s disease (AD), Huntington Disease, or progressive supranuclear palsy (PSP) ^62-68^.

Intriguingly, recent studies by Schellenberg and colleagues raise the possibility that variants of *EIF2AK3*, which encodes PERK, may also be a genetic risk factor in neurodegenerative diseases based on the finding that a specific protein-coding haplotype of *EIF2AK3* was associated with increased risk for PSP through genome-wide association and postmortem studies ^69; 70^. To date, three major coding haplotypes of *EIF2AK3* have been identified; of these, the common haplotypes are A (64%) and B (29%) based on 26 genome-wide association studies (GWASs) (http://www.ncbi.nlm.nih.gov/SNV/), whereas the less common D haplotype has a frequency of 6% in the 1000 Genome data (Table 1). Schellenberg *et al*. showed that *EIF2AK3* haplotype B harboring minor alleles of three nonsynonymous single nucleotide variants (SNVs), including rs7571971, in the coding region was associated with increased PSP risk ^69; 70^. The authors also reported that the rs7571971 risk allele was in strong linkage disequilibrium with the haplotype B-coding SNVs. Another study demonstrated that lymphoblastoid cell lines derived from *EIF2AK3* haplotype B-expressing individuals had enhanced PERK activity compared to those lines derived from *EIF2AK3* haplotype A-expressing individuals ^71^; furthermore, PERK inhibition by small-molecule inhibitors or by modulation of eIF2α phosphorylation was reported to alleviate neurological deficits and pathology in small animal models of several neurodegenerative diseases ^72-79^.

**Table 1.** Major haplotypes of *EIF2AK3*

|  | **rs867529-rs13045-rs1805165** | **Amino acids** |
| --- | --- | --- |
| **Haplotype A** | CGT | Ser136-Arg166-Ser704 |
| **Haplotype B** | GAG | Cys136-Gln166-Ala704 |
| **Haplotype D** | CAT | Ser136-Gln166-Ser704 |

In light of the evidence showing increased or sustained PERK activity as detrimental to several critical cellular pathways in a wide range of conditions and based on the possibility of a link between specific minor alleles of *EIF2AK3* and neurodegenerative diseases with features shared by NCI in PWH, we hypothesized that minor *EIF2AK3* alleles such as haplotype B-associated SNVs are a risk factor for NCI in PWH. The objective of the present study was to determine whether minor alleles of *EIF2AK3* SNVs, which were previously reported to be associated with neurodegeneration, correlated with worse NC performance in PWH.

## Materials and Methods

### Study cohort

This was a retrospective study of data prospectively collected from 1,047 PWH who were assessed in the CNS HIV Anti-Retroviral Therapy Effects Research (CHARTER) cohort ^2^ between 2003 and 2015 and who had genomic DNA available for analysis. Participants who had severe neuropsychiatric comorbidities (e.g., untreated schizophrenia or seizure disorder) were excluded. All participants were comprehensively evaluated with neuromedical and NC assessments at least once, and 566 participants were assessed at least twice (median 7 assessments, interquartile range 4-11). Blood was collected by venipuncture and cerebrospinal fluid (CSF) was collected by lumbar puncture. All study procedures, including genetic testing, were approved by the University of California San Diego Human Research Protections Program, and all participants provided written informed consent for the study procedures, including future use of data, biospecimens, and genetic data.

Genomic data were analyzed in two phases. The first phase included the genome-wide data of1,047 participants in the CHARTER cohort. The data were queried for three noncoding SNVs in *EIF2AK3* (rs6739095, rs1913671, rs11684404), which were previously reported to be in linkage disequilibrium with the three coding SNVs in *EIF2AK3*. In parallel, targeted sequencing (TS) was performed in 992 of the 1,047 individuals with available genomic DNA to determine the genotypes of three coding SNVs in *EIF2AK3* (rs867529, rs1805165, and rs13045), which determine the A, B, and D haplotypes of *EIF2AK3*.

### Targeted sequencing

Genomic DNA samples of the 992 individuals were analyzed using TaqMan SNV genotyping assays (Life Technologies) for rs867529, rs1805165, and rs13045. The assays were performed by polymerase chain reaction according to the previously published conditions ^70^. Genotypes were visualized and called using a 7900HT Fast Real Time PCR system and the allelic discrimination function of the Sequence Detection System V.2.4 (Applied Biosystems, Waltham, MA, USA).

### Neuropsychiatric Assessment

All participants completed standardized, comprehensive neuropsychological assessments of seven cognitive domains that are commonly affected by HIV (learning, recall, motor function, executive function, working memory, speed of information processing, and verbal fluency) ^80^. The best available normative standards corrected for the effects of age, education, sex, and race/ethnicity. Test scores were converted to demographically corrected standard T scores, which were then converted into deficit scores. The deficit scores across the entre battery were then averaged to derive a Global Deficit Score (GDS), which ranges from 0 to 5, with higher scores indicating poorer NC performance ^81^. A GDS value ≥ 0.5 indicates NCI. Depressive symptoms were assessed by the Beck Depression Inventory Second Edition (BDI-II), a validated survey of 21 questions that assess the intensity, severity and depth of depressive symptoms.^82^ Higher BDI-II values indicate more depressive symptoms with a value > 13 indicating at least mild depression.

### Statistical Analysis

To assess explanatory variables for impaired vs unimpaired participants in the cohorts, categorical data were compared using Pearson’s chi-square test, and continuous data were compared using Student’s *t* test. Univariable and multivariable methods, including logistic and linear regression, were used to determine the association of the noncoding and coding SNVs with demographic characteristics (e.g., age, sex, and ancestry), disease-related variables (e.g., plasma and cerebrospinal fluid (CSF) HIV RNA, nadir CD4^+^ T cell count, comorbidity (e.g., depression, HCV coinfection), treatment status (e.g., ART use), and NC performance, including global deficit score (GDS), global NCI, and domain-specific deficit scores (DDSs). To account for type I error in the analyses of multiple variables associated with GDS and NCI, the false discovery rate approach was used. Akaike Information Criterion and backward selection were used in multivariable regression. All statistical analyses were performed using JMP Pro version 15.0 (SAS Institute, Cary, NC, USA).

## Results

### The characteristics of the participants

Table 2 summarizes the characteristics of the 1,047 individuals who had data on noncoding *EIF2AK3* SNVs. Briefly, the mean age was 43.1 years, 22.8% of the cohort were female, and 41% had European ancestry. Most participants had AIDS (60.7%) and were categorized as having only “minimal” neuropsychiatric comorbidities (64.1%). The median CD4^+^ T cell counts were 175/µL (nadir) and 428/µL (current). At the first assessment, 70.5% used ART and 68.3% of these had plasma HIV RNA levels ≤ 200 copies/mL.

**Table 2.** Characteristics of the participants according to the neurocognitive impairment status

|  | All<br>(n = 1047) | Unimpaired<br>(n = 663) | Impaired<br>(n = 384) | *p* |
| --- | --- | --- | --- | --- |
| Age, y, mean | 43.1 | 43.0 | 43.4 | 0.26 |
| Sex, female | 22.8% | 24.2% | 22.1% | 0.456 |
| Ancestry |  |  |  | **0.002** |
| European ancestry | 41.0% | 40.8% | 41.4% | 0.84 |
| Admixed Hispanic | 12.3% | 9.9% | 17.0% | **0.001** |
| African ancestry | 46.7% | 49.3% | 41.7% | **0.003** |
| NP comorbidity | 64.1% | 70.4% | 53.1% | **<0.001** |
| AIDS diagnosis | 60.7% | 58.2% | 65.6% | **0.021** |
| HCV seropositivity | 25.9% | 26.1% | 24.2% | 0.36 |
| Nadir CD4<sup>+</sup> T cell<br>count (/μL), median<br>(IQR) | 175 (50-<br>300) | 190 (54-320) | 150 (36-257) | **0.028** |
| Current CD4<sup>+</sup> T<br>Cells (/μL), median<br>(IQR) (n = 1040) | 428 (265–<br>600) | 426 (269–593) | 434 (250–607) | 0.97 |
| ART – current use | 70.5% | 67.4% | 76.4% | **0.002** |
| Plasma HIV RNA ≤<br>200 copies/mL (on<br>ART) | 68.3% | 69.2% | 66.3% | 0.41 |

AIDS, acquired immunodeficiency syndrome; ART, antiretroviral therapy; GWAS, genome-wide association study; HCV, hepatitis C virus; HIV, human immunodeficiency virus; IQR, interquartile range; NP, neuropsychiatric

Consistent with prior reports from CHARTER, analyses identified that NCI (n=384, 37.4%) was associated with AIDS (65.6% vs 58.2% in the 663 participants without NCI, *p* = 0.021), ART use (76.4% vs 67.4%, *p* = 0.002), and lower nadir CD4^+^ T cell count (150 vs 190/µL, *p* = 0.028), and more than minimal comorbidities (70.4% vs 53.1%, *p* < 0.001).

Regarding the noncoding SNVs, 41.7% of the cohort had at least one T minor allele for rs6739095, 41.4% of the cohort had at least one C minor allele for rs1913671, and 39.4% of the cohort had at least one C minor allele for rs11684404. The frequencies of the minor alleles of the three noncoding SNVs reflected those reported in the general population (Table S1). The concordance among the three minor alleles of the three noncoding SNVs was also high in the CHARTER cohort (Table S2), in agreement with previous GWAS findings.

### Minor alleles of noncoding *EIF2AK3* SNVs are associated with worse GDS and NCI

We first examined the association of noncoding *EIF2AK3* SNVs with GDS in the GWAS dataset. As shown in Figure 1, we found that the minor alleles of all three noncoding SNVs were associated with worse GDS and NCI in a dose-dependent manner. For example, the mean GDS values were 0.43 (TT), 0.50 (CT), and 0.56 (CC) for the rs11684404 C risk allele (Fig. 1A, *p* = 0.0016). The frequency of NCI was also significantly higher in CC homozygotes than in TT homozygotes for rs11684404 (Fig. 1D, 46.3% vs 32.3%, *p* = 0.0262). Similarly, the rs6739095 T and rs1913671 C risk alleles were associated with GDS as well as with NCI (Fig. 1B, 1C, 1E, 1F).

**Figure 1.**
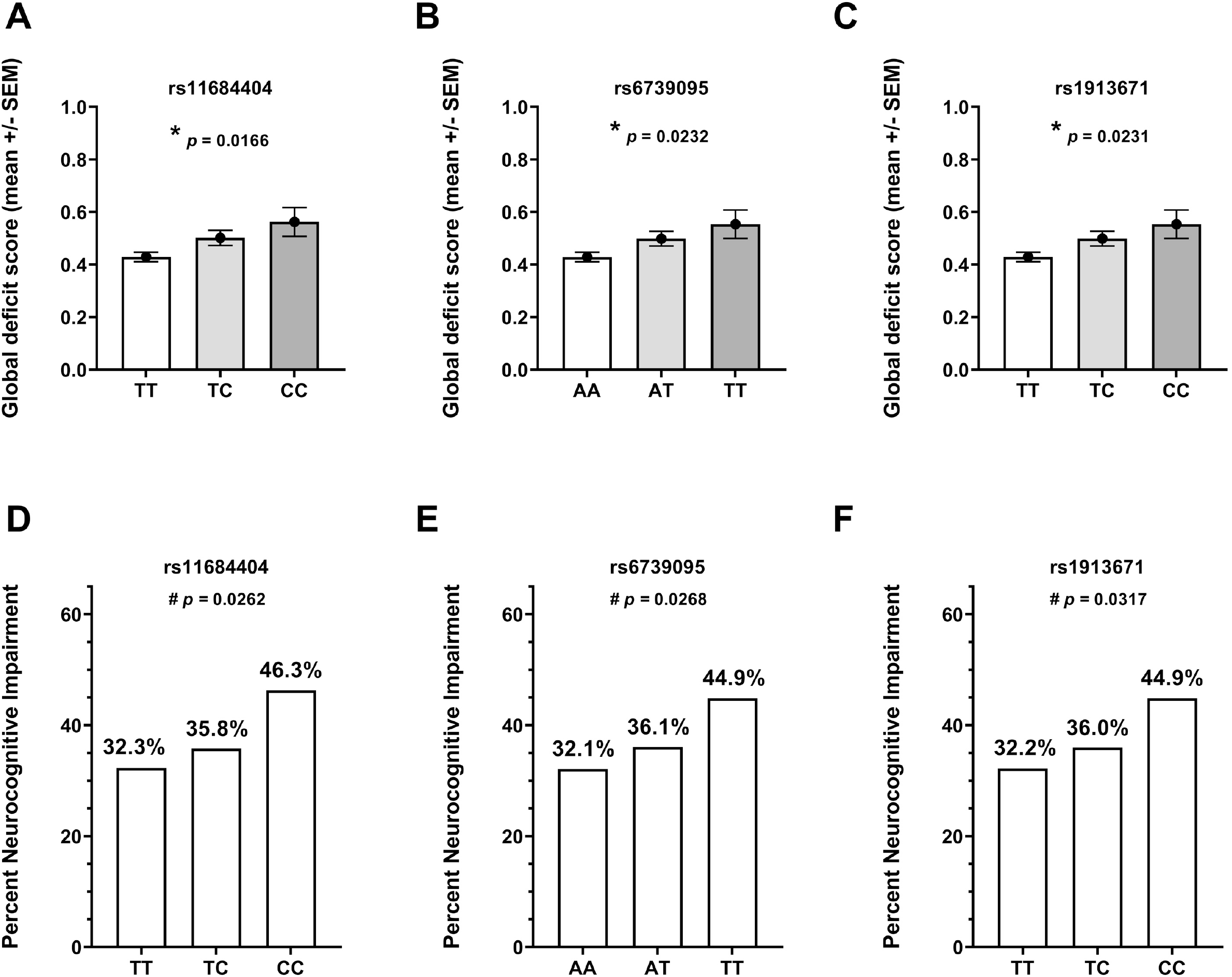
Association of noncoding *EIF2AK3* SNVs with GDS (A–C) and NCI (D–E) in the GWAS cohort GDS, global deficit score; NIC, neurocognitive impairment; SNV, single nucleotide variant * *p* < 0.05, analysis of variance; ^#^ *p* < 0.05, Cochran–Armitage test

### Minor alleles of noncoding *EIF2AK3* SNVs are associated with performance in specific NC ability domains

Emerging data from recent studies including CHARTER and the MACS/WIHS Combined Cohort Study (MWCCS) support that several indicators such as sex and inflammation are associated with deficits in specific NC domains, such as delayed recall or working memory ^11; 83-87^. We analyzed the associations of the three noncoding *EIF2AK3* SNVs with deficits in NC domains in the CHARTER GWAS dataset. As shown in Table 3, the minor alleles of all three noncoding SNVs were associated with deficits in executive functioning (*p* < 0.001), verbal fluency (*p* < 0.05), motor function (*p* < 0.05), and possibly learning (p values, 0.038 to 0.054).

**Table 3.** Association of noncoding *EIF2AK3* SNVs with domain-specific scores

| Domain-specific T score | *p* value |  |  |
| --- | --- | --- | --- |
|  | rs6739095 | rs1913671 | rs11684404 |
| Verbal | **0.0256** | **0.0198** | **0.0103** |
| Executive | **<0.001** | **<0.001** | **<0.001** |
| Motor | **0.0117** | **0.0230** | **0.0138** |
| Learning | **0.0542** | **0.0524** | **0.0381** |
| Recall | 0.7697 | 0.7779 | 0.9020 |
| Working memory | 0.3534 | 0.3962 | 0.4429 |
| Speed of information processing | 0.354 | 0.350 | 0.336 |

SNV, single nucleotide variant

### Minor alleles of noncoding *EIF2AK3* SNVs are independently associated with GDS and NCI

Multiple linear regression analysis was conducted to explore the association of the noncoding *EIF2AK3* SNVs and the HIV and host factors previously shown to be related to GDS. The analysis revealed that the minor allele of rs11684404 remained significantly associated with GDS, along with ART use and neuropsychiatric comorbidities (*R*^*2*^ = 0.08, *p* < 0.0001) (Table 4), whereas the minor alleles of the other two noncoding *EIF2AK3* SNVs did not remain significantly associated with GDS after accounting for the influence of rs11684404. In contrast, multiple logistic regression analysis identified that NCI was associated with the minor allele of rs6739095, but not rs11684404, as well as with ART use, neuropsychiatric comorbidities, and HCV seropositivity (*R*^*2*^ = 0.04, *p* < 0.0001) (Table 4).

**Table 4.** Multivariable analysis of the CHARTER dataset

|  | Univariable |  | Multivariable |  |
| --- | --- | --- | --- | --- |
| | $\beta$ | *p* value | $\beta$ | *p* value |
| **GDS (Model $R^2=0.080$, $p=2.1 \times 10^{-16}$)** |  |  |  |  |
| rs6739095, T | 0.08 | **0.0058** |  |  |
| rs11684404, C | 0.083 | **0.0026** | 0.048 | **0.0017** |
| Neuropsychiatric Comorbidities | 0.111 | **<0.001** | 0.111 | **<0.001** |
| ART Use | 0.058 | **<0.001** | 0.038 | **0.047** |
| Nadir CD4<sup>+</sup> T-Cells | $-2.5 \times 10^{-4}$ | **0.0018** | $-1.7 \times 10^{-4}$ | **0.059** |
| AIDS diagnosis | 0.042 | **0.006** |  |  |
| BDI-II > 13 | 0.041 | **0.012** | 0.023 | **0.153** |
| HIV RNA log<sub>10</sub> c/mL | -0.027 | **0.021** |  |  |
| Age | 0.003 | **0.077** |  |  |
| Ancestry | -0.026 | **0.100** | -0.027 | **0.085** |
| **NCI (Model $R^2=0.044$, $p=1.1 \times 10^{-11}$)** |  |  |  |  |
| | $\beta$ | *p* value | $\beta$ | *p* value |
| rs6739095, T | 0.285 | **0.024** | 0.164 | **0.015** |
| rs11684404, C | 0.299 | **0.018** |  |  |
| Neuropsychiatric Comorbidities | 0.371 | **<0.001** | 0.401 | **<0.001** |
| ART Use | 0.267 | **<0.001** | 0.279 | **<0.001** |
| Nadir CD4<sup>+</sup> T-Cells | 0.001 | **0.0046** |  |  |
| BDI-II > 13 | 0.163 | **0.018** | 0.104 | **0.148** |
| AIDS diagnosis | 0.142 | **0.032** |  |  |
| HCV serostatus | -0.126 | **0.096** | -0.221 | **0.005** |

Candidate covariates tested included demographic (e.g., age, race, sex), HIV disease (e.g., HIV RNA in plasma, nadir and current CD4<sup>+</sup> T cell counts), HIV treatment (e.g., ART use), and common comorbidities (e.g., neuropsychiatric comorbidities, HCV co-infection). The *p* values for BDI-II > 13 are greater than 0.05 but the covariate was retained in the model by Akaike Information Criterion selection.
AIDS, acquired immunodeficiency syndrome; ART, antiretroviral therapy; BDI-II, Beck Depression Inventory-II; GWAS, genome-wide association study; HCV, hepatitis C virus; HIV, human immunodeficiency virus; IQR, interquartile range; NP, neuropsychiatric

### Minor alleles of coding *EIF2AK3* SNVs are associated with worse GDS and NCI

Based on our analyses indicating associations of noncoding *EIF2AK3* SNVs with GDS and NCI, we next analyzed the minor alleles of three coding *EIF2AK3* SNVs, which were previously shown to be associated with increased risk of PSP^69; 70^. The characteristics of this TS sub-cohort, which included 992 participants with available genomic DNA for analysis, are summarized in Table S3. Risk alleles were again present in a substantial minority: at least one A allele for rs13045 in 41.3%, at least one G allele for rs867529 in 30.9%, and at least one G allele for rs1805165 in 30.9%. The frequencies of the minor alleles of these three SNVs are consistent with their reported frequencies across the 26 published GWASs (Table S4). However, the concordance among the three coding SNVs was different than that observed among the three noncoding SNVs. Specifically, rs867529 and rs1805165 were 100% concordant whereas each exhibited 88.3% concordance with rs13045 (Table S5). Even with this difference in concordance, the minor alleles of all three coding SNVs were also significantly associated with GDS and NCI in a dose-dependent manner, as shown in Figure 2.

**Figure 2.**
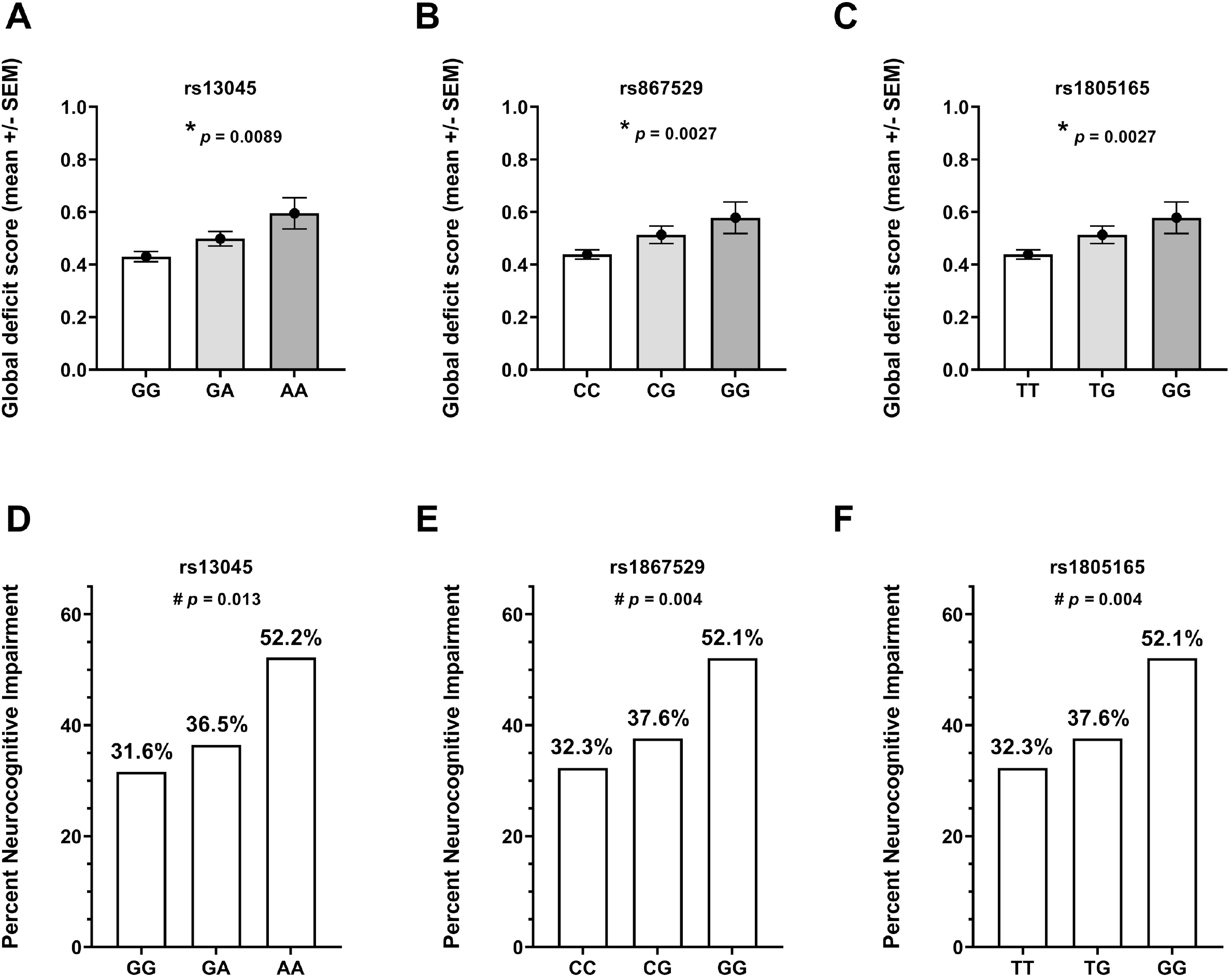
Association of coding *EIF2AK3* SNVs with GDS (A–C) and NCI (D–E) in the TS cohort * *p* < 0.05, analysis of variance; ^#^ *p* < 0.05, Cochran–Armitage test

### Minor alleles of coding *EIF2AK3* SNVs are associated with deficits in cognitive domains

Our analyses to investigate the associations of the three coding *EIF2AK3* SNVs with deficits in specific NC domains in the TS cohort revealed distinct patterns, as shown in Table 5. Specifically, the minor allele of rs13045 was associated with domain-specific deficits in executive functioning (*p* = 0.0131), learning (*p* = 0.0213), and motor domains (*p* = 0.0116) whereas the minor alleles of rs867529 and rs1805165 were significantly associated with domain-specific deficits in verbal fluency (*p* = 0.0025) and motor domains (*p* = 0.0078).

**Table 5.** Association of coding *EIF2AK3* SNVs with domain-specific scores

| Domain-specific T score | *p* value |  |  |
| --- | --- | --- | --- |
|  | rs13045 | rs867529 | rs1805165 |
| Motor | **0.0116** | **0.0078** | **0.0078** |
| Verbal | **0.081** | **0.0025** | **0.0025** |
| Executive | **0.0131** | **0.0518** | **0.0518** |
| Learning | **0.0213** | 0.1259 | 0.1259 |
| Recall | 0.1368 | 0.0650 | 0.0650 |
| Working memory | 0.5629 | 0.1032 | 0.1032 |
| Speed of information processing | 0.229 | 0.358 | 0.358 |

SNV, single nucleotide variant

### Minor alleles of coding *EIF2AK3* SNVs are associated with increased CSF levels of IL-6

Based on studies finding that IL-6 concentrations in blood and CSF are a biomarker of inflammation that is associated with NCI in PWH ^88^, we assessed the relationship of the coding *EIF2AK3* SNVs with this biomarker in participants with CSF IL-6 concentrations that were previously quantified by immunoassay and stored in the CHARTER data repository (n = 534). The CSF IL-6 levels did not significantly differ between those with and without NC impairment (Fig. 3A). The minor alleles of all three coding *EIF2AK3* SNVs were significantly associated with higher IL-6 in CSF (Figure 3B, 3C; rs13045: *p* = 0.024, rs867529: *p* = 0.011, and rs1805165: *p* = 0.011; data not shown for rs1805165). In contrast, the minor alleles of these coding SNVs were not associated with IL-6 concentration in blood plasma among participants with available data (n = 144; data not shown).

**Figure 3.**
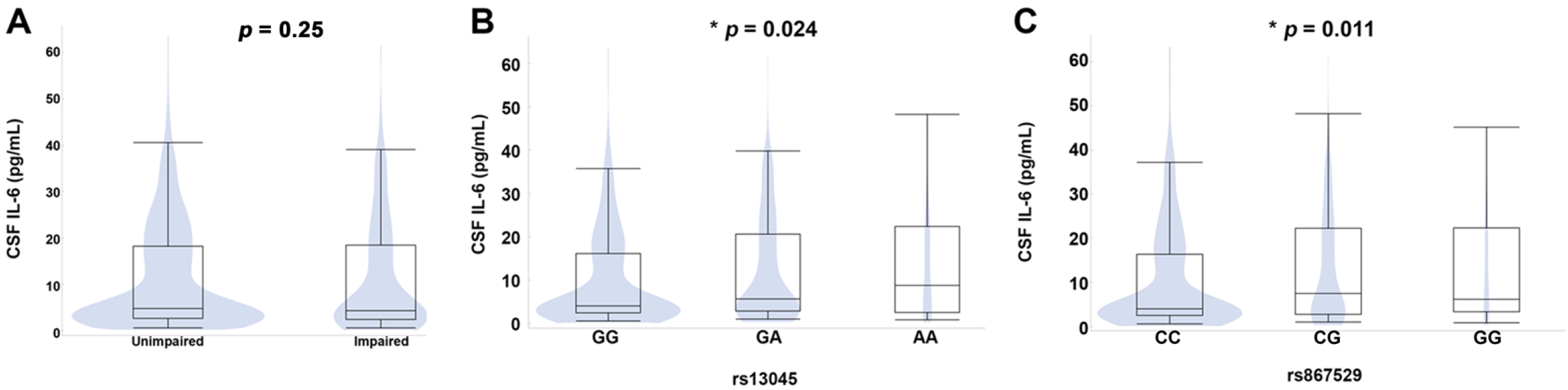
Association of NCI (A) and two coding *EIF2AK3* SNVs (B, C) with CSF IL-6 levels in a subset of the study cohort with available data * *p* < 0.05, analysis of variance

### Minor allele of rs13045 is independently associated with GDS and NCI

Our multiple logistic regression analysis comparing the coding *EIF2AK3* SNVs to HIV and host factors revealed that the minor allele of rs13045 remained significantly associated with NCI, along with ART use and a BDI-II score >13 ^82; 89^ (*R*^*2*^ = 0.048, *p* < 0.0001) (Table 6), although the minor alleles of the other two coding *EIF2AK3* SNVs did not. Multiple linear regression of GDS confirmed this finding: only the rs13045 minor allele (but not the other coding SNVs) remained associated with GDS, along with nadir CD^+^ T-cell count, BDI-II > 13, and European ancestry (*R*^*2*^ = 0.03, *p* < 0.0001) (Table 6).

**Table 6.** Multivariable analysis of the targeted sequencing sub-cohort for NCI and GDS

| **GDS (Model R<sup>2</sup>=0.03, p < 0.0001)** |  |  |  |  |  |
| --- | --- | --- | --- | --- | --- |
|  | **Risk direction** | **β** | **p value** | **β** | **p value** |
| rs13045 | A allele | 0.076 | **0.002** | **0.057** | **0.025** |
| Neuropsychiatric comorbidities | Yes | 0.074 | **0.017** |  |  |
| ART Use | Current use | 0.072 | **0.041** |  |  |
| Nadir CD4<sup>+</sup> T-Cells | Lower | $-1.7 \times 10^{-4}$ | **0.045** | $-1.7 \times 10^{-4}$ | **0.048** |
| AIDS diagnosis | Yes | 0.076 | **0.016** |  |  |
| BDI-II > 13 | >13 | 0.107 | **<0.001** | **0.096** | **0.002** |
| HIV RNA log<sub>10</sub> c/mL | Lower | -0.022 | 0.07 |  |  |
| Age | Older | 0.003 | 0.141 |  |  |
| Ancestry | European | 0.111 | **<0.001** | **0.093** | **0.004** |
| **NCI (Model R<sup>2</sup>=0.048, p < 0.0001)** |  |  |  |  |  |
|  | **Risk direction** | **β** | **p value** | **β** | **p value** |
| rs13045 | A allele | 0.338 | **0.015** | 0.339 | **0.003** |
| Neuropsychiatric comorbidities | Yes | 0.28 | **0.041** |  |  |
| ART Use | Current use | 0.530 | **0.001** | 0.504 | **0.002** |
| Nadir CD4<sup>+</sup> T-Cells | Lower | $-6 \times 10^{-4}$ | 0.14 | | |
| AIDS diagnosis | Yes | 0.19 | 0.17 |  |  |
| BDI-II > 13 | >13 | 0.539 | **<0.001** | 0.534 | **<0.001** |
| HIV RNA log<sub>10</sub> c/mL | Lower | -0.117 | **0.031** |  |  |
| Age | Older | 0.006 | 0.44 |  |  |
| Ancestry | European | 0.342 | **0.012** |  |  |

ART, antiretroviral therapy; BDI-II, Beck Depression Inventory-II; GDS, global deficit score; NCI, neurocognitive impairment

### Association of *EIF2AK3* haplotypes with NCI and GDS

Based on the complete concordance between rs867529 and rs1805165, we constructed *EIF2AK3* haplotypes in the TS cohort. The frequency of these haplotypes were *EIF2AK3* A/A 58.7% (n = 583), A/B 24.7% (n = 245), A/D 9.8% (n = 97), B/B 4.8% (n = 48), B/D 1.4% (n = 14), and D/D 0.5% (n = 5) (Table S6). The haplotype groups did not differ in participant characteristics. *EIF2AK3* haplotypes were not associated with NCI or GDS (data not shown) or with IL-6 concentrations in CSF (Figure S1).

## Discussion

One of the conundrums of the ART era is the persistence of NCI even in PWH who have achieved viral suppression and immune recovery with ART. Up to 32.7% of PWH exhibit NCI-related dysfunction in activities of daily living and even asymptomatic NCI in PWH confers substantial risk for transitioning to symptomatic status over a period of just a few years ^2-6; 90^. In this retrospective analysis of the well characterized CHARTER cohort, we found that previously defined coding and noncoding *EIF2AK3* minor risk alleles were significantly associated with global NCI and the continuous GDS impairment measure. We also observed that the minor alleles of all three coding *EIF2AK3* SNVs were associated with higher IL-6 concentrations in CSF. Furthermore, the associations between the rs13045 A minor risk allele and either global NCI or GDS remained significant after adjustment for other covariates in multivariable models. The three coding *EIF2AK3* minor risk alleles were associated with performance in four NC domains: motor functioning, verbal fluency, executive functioning, and learning. While recent studies have reported certain coding and noncoding *EIF2AK3* SNVs as genetic risk factors in several neurodegenerative diseases, including AD and PSP ^69; 70; 91^, this is the first study to demonstrate an association between multiple coding and noncoding *EIF2AK3* SNVs and cognition in PWH. Overall, these findings support accumulating evidence of ISR activation as a contributor to the pathogenesis of HIV in the CNS ^62-64; 92-98^ and implicate *EIF2AK3* as a potentially important component of genetic vulnerability to NCI in PWH.

Among the four major *EIF2AK3* haplotypes, haplotypes A and B encode protein products that differ by changes in two amino acids in the luminal domain (S136C, R166Q) and one amino acid in the kinase domain (S704A) of PERK (Table 1). The significant associations of minor risk alleles of *EIF2AK3* coding SNVs with NC performance uncovered in the present study are likely related to differences in PERK activation, activity, or downregulation resulting from these amino acid changes. However, the impact of these *EIF2AK3* SNVs on PERK kinase activity and downstream cellular events remains unclear due to contradictory findings reported by a limited number of studies. In one study, lymphoblastoid cell lines derived from *EIF2AK3* haplotype B-expressing individuals, which exhibited similar baseline PERK protein levels, nonetheless displayed enhanced PERK kinase activity, as determined by peIF2α accumulation, in response to challenge with tunicamycin, which blocks N-linked glycosylation and induces ER stress ^71^. In contrast, another study proposed that *EIF2AK3* haplotype B might be a hypomorph based on higher eIF2α phosphorylation in *EIF2AK3* haplotype A-expressing cells compared to *EIF2AK3* haplotype B-expressing cells, both at baseline and following treatment with thapsigargin, which inhibits the sarcoplasmic/ER Ca2^+^ ATPase pump ^99^. The association of specific *EIF2AK3* SNVs with increased kinase activity and deleterious outcomes is likely cell- and context-dependent. Nonetheless, our analyses showing the significant association of the minor alleles of the three coding *EIF2AK3* SNVs with NC performance corroborate our previous studies demonstrating ISR activation in the post-mortem brain tissue from ART-treated PWH ^62; 63^ and implicate that maladaptive signaling might be at play in the CNS of PWH harboring the minor alleles of *EIF2AK3* SNVs.

In the TS analysis, only the rs13045 A allele remained significantly associated with both GDS and NCI in multivariable analyses. The rs13045 minor allele is present in both haplotypes B and D, whereas haplotype D is defined by the presence of the rs13045 A allele alone. The frequency of the rs13045 A allele is higher than the frequencies of the other two coding *EIF2AK3* SNVs across different racial groups around the globe (Table S7), which might partially explain the lack of association of the other two SNVs in multivariable analysis in the present study. The number of haplotype D heterozygotes or homozygotes was small (116/992), precluding more in-depth analysis. In addition to the need for follow-up studies in larger cohorts to clarify whether haplotype D confers increased risk for NCI, mechanistic studies are also warranted to determine whether the change in amino acid 166 from arginine to glutamine in cells carrying the rs13045 A allele may facilitate weaker affinity for binding immunoglobulin protein (BiP), allowing its dissociation from PERK, with subsequent reduction in PERK kinase activation threshold.

Our multivariable analyses of the TS cohort also confirmed that a level of depressive symptoms consistent with at least mild depression (BDI-II > 13), was also significantly associated with NC performance, supporting the contribution of depression to worse NC outcomes in PWH, which that has been recently reported in the CHARTER cohort ^100; 101^. These results also implicate a potential interaction between the rs13045 A allele and depression, which is currently under investigation. As the most common neuropsychiatric comorbidity in PWH ^7; 8^, depression is associated with worse ART adherence, viral suppression ^102; 103^, and survival ^104; 105^. Although no study to date has reported a link between *EIF2AK3* SNVs and depression, the expression levels of several UPR-related genes and proteins, including *EIF2AK3* and PERK, are higher in adults with major depressive disorder ^106; 107^. In addition, depressive-like behaviors and memory impairment can be alleviated by inhibiting PERK-peIF2α signaling in animal models of depression ^108-112^.

The observed association between an *EIF2AK3* SNV and higher IL-6 concentrations in CSF is particularly interesting since several studies have linked systemic and CNS inflammation to neuropsychiatric conditions, including worse NC performance and depressive symptoms ^89; 113^. This finding provides additional evidence for a potential role of PERK in neuroinflammation in several disorders including NCI in PWH ^94; 114; 115^.

The significant association of the minor alleles of specific *EIF2AK3* SNVs with deficits in specific domains raise the possibility of a link between specific SNVs and certain NC phenotypes, including declines in verbal fluency, executive function, learning, and motor function, which have been recently described in the MWCCS ^86; 116; 117^. These phenotypes likely differ in viral, host, and genetic mechanisms. Intriguingly, Kolson and colleagues recently reported associations between regional brain neuroinflammation and heme oxygenase-1 (HO-1) responses in analyses that included brain tissue from autopsy and CSF from PWH taking ART ^118; 119^. Their findings also implicate regional HO-1 isoform expression may be associated with differences in recovery from acute synaptic injury in a primate model of lentiviral infection ^120^. Given the well-characterized mechanistic role of PERK and HO-1 in activating the antioxidant response ^39^, an interaction between *EIF2AK3* SNVs and regional susceptibility to oxidative stress might partially contribute to the observed associations of specific minor alleles of *EIF2AK3* SNVs with NC phenotypes.

The construction of the *EIF2AK3* haplotypes in the TS cohort revealed that the frequencies of specific haplotypes were similar to those reported in the general population, supporting that our findings may be generalizable (Table S6). The comparison of domain-specific deficit scores among groups categorized according to haplotypes suggest that certain haplotypes might be associated with deficits in certain domains (Table S8); however, the number of participants in certain groups was low, precluding further statistical analyses. Future studies are warranted in other, larger cohorts such as the MWCCS, which has notable differences including greater inclusion of women and people without HIV as well as a different distribution of ancestry, to determine the potential role of *EIF2AK3* as a genetic risk factor for NCI in PWH.

Our retrospective analysis of the CHARTER GWAS data revealed that the minor alleles of the three noncoding *EIF2AK3* SNVs, which were in strong linkage disequilibrium with those of the three coding *EIF2AK3* SNVs, were also significantly associated with GDS and NCI. The impact of these noncoding SNVs on PERK expression, function or stability is unclear, although they have previously been linked to increased risk for PSP ^70^. All three noncoding SNVs are located in the intronic regions of *EIF2AK3* and may result in the formation of alternative transcripts leading to RNA species or protein isoforms with altered stability or function, which warrants further investigation.

In light of the evidence showing increased or sustained PERK activity as detrimental to several critical cellular pathways in a wide range of conditions ^55; 56; 59; 61; 72; 74; 75^, we propose that minor alleles of SNVs associated with increased *EIF2AK3* activity in response to stress might be a predictive risk factor for NCI in PWH. Aging-related comorbid diseases may cause sustained ISR activation via PERK ^64; 72; 92; 95; 121; 122^ that may further increase the risk for NCI in PWH.

Our findings raise several fundamental questions that should be addressed before establishing *EIF2AK3* as a risk factor for NCI in PWH. First, the regulation and kinase activity of disease-associated *EIF2AK3* SNVs should be described. Second, mechanisms underlying the association of specific *EIF2AK3* SNVs with deleterious cellular responses to disease-relevant stressors should be elucidated in pertinent cellular contexts. Furthermore, whether the observed association of these SNVs with specific domains can be replicated and used to reliably identify specific NC phenotypes should be clarified. Future studies should also explore the relationship of *EIF2AK3* SNVs with molecular and imaging biomarkers of NCI in PWH. These studies should assess their contribution to a polygenic risk score as part of a precision medicine approach to identify phenotypes and specific populations who may specifically benefit from adjunctive therapies such as modulators of PERK and endogenous antioxidant response during the best therapeutic window, soon after HIV diagnosis.

The present study has several limitations that should be addressed in future studies. First, the study cohort was relatively small for a genetics study. However, the distributions of both noncoding and coding *EIF2AK3* SNVs in the CHARTER cohort were similar to those reported in larger GWASs ^70^. Additionally, the concordance among the noncoding and coding *EIF2AK3* SNVs followed the expected pattern reported in the general population. Second, due to the low number of cases with some haplotypes, we were unable to examine the relationship of *EIF2AK3* haplotypes with CSF IL-6 concentrations or BDI-II scores. The cross-sectional design precludes the inference of a causal relationship between *EIF2AK3* haplotypes and NC impairment. Studies in animal models are warranted to reveal the mechanistic underpinnings of the observed associations. Finally, the models did not explain substantial variance in GDS or NCI. While this is true, the effect size for *EIF2AK3* SNVs was similar in magnitude to effect sizes attributable to ART use, neuropsychiatric comorbidities, and other covariates, highlighting the potential importance of this genetic vulnerability.

In summary, the observed associations of minor alleles of *EIF2AK3* SNVs with NCI, albeit small-to-medium in size, were present in multivariable analyses, raising the possibility of specific SNVs in *EIF2AK3*, and perhaps other ISR-related genes, as an important component of genetic vulnerability in PWH. Determination of host factors that are predictive for NC phenotypes early in the course of HIV disease might enable more effective preventive and therapeutic interventions for NC disorders.

## Data Availability

All data produced in the present study are available upon reasonable request to the authors.

## Supplemental Data

Supplemental data include one figure and eight tables.

## Declaration of interests

All authors declare that they have no relevant disclosures or conflicts of interest.

## Acknowledgments

The CNS HIV Anti-Retroviral Therapy Effects Research was supported by awards N01 MH22005, HHSN271201000036C, HHSN271201000030C, and R01 MH107345 from the National Institutes of Health.

The CNS HIV Anti-Retroviral Therapy Effects Research (CHARTER) group is affiliated with Johns Hopkins University; the Icahn School of Medicine at Mount Sinai; University of California, San Diego; University of Texas, Galveston; University of Washington, Seattle; Washington University, St. Louis; and is headquartered at the University of California, San Diego and includes: Directors: Robert K. Heaton, Ph.D., Scott L. Letendre, M.D.; Center Manager: Donald Franklin, Jr.; Coordinating Center: Brookie Best, Pharm.D., Debra Cookson, M.P.H, Clint Cushman, Matthew Dawson, Ronald J. Ellis, M.D., Ph.D., Christine Fennema Notestine, Ph.D., Sara Gianella Weibel, M.D., Igor Grant, M.D., Thomas D. Marcotte, Ph.D. Jennifer Marquie-Beck, M.P.H., Florin Vaida, Ph.D.; Johns Hopkins University Site: Ned Sacktor, M.D. (P.I.), Vincent Rogalski; Icahn School of Medicine at Mount Sinai Site: Susan Morgello, M.D. (P.I.), Letty Mintz, N.P.; University of California, San Diego Site: J. Allen McCutchan, M.D. (P.I.); University of Washington, Seattle Site: Ann Collier, M.D. (Co-P.I.) and Christina Marra, M.D. (Co-P.I.), Sher Storey, PA-C.; University of Texas, Galveston Site: Benjamin Gelman, M.D., Ph.D. (P.I.), Eleanor Head, R.N., B.S.N.; and Washington University, St. Louis Site: David Clifford, M.D. (P.I.), Mengesha Teshome, M.D.

The views expressed in this article are those of the authors and do not reflect the official policy or position of the United States Government.

## Funding

This project was supported by the following grants: R01 MH126773 and R01 MH098742 (KJS), R01 MH109382 (KJS, CAE), UG3NS104095 NINDS (GDS); N01 MH22005, HHSN271201000036C, HHSN271201000030C, and R01 MH107345 (CHARTER Study: SLL, RJE, RKH, IG, DRF); R01MH095621 (AK).

## Authors’ contributions

CAE performed targeted sequencing and analyses of coding SNVs and haplotypes, wrote the first draft of the manuscript. SB provided intellectual feedback and edited the manuscript. BD performed targeted sequencing. AK provided intellectual feedback, and edited the manuscript. AB provided intellectual feedback and edited the manuscript. DRF provided intellectual feedback and edited the manuscript. GDS provided intellectual feedback and edited the manuscript. RKH provided intellectual feedback and edited the manuscript. IG served as PI of CHARTER and was involved in design, obtaining funding, and intellectual contribution. SLL and RJE designed, obtained funding for, and conducted the CHARTER project and edited the manuscript. SLE performed analyses of noncoding SNVs and his lab also performed the IL-6 assays. KJS obtained funding for the study, performed analyses of coding SNV and haplotypes, and edited the manuscript.

## Notes

### Competing Interest Statement

The authors have declared no competing interest.

### Author Declarations

All study procedures, including genetic testing, were approved by the University of California San Diego Human Research Protections Program, and all participants provided written informed consent for the study procedures, including future use of data, biospecimens, and genetic data.

