## Supplementary figures and images for "Genetic variations in *EIF2AK3* are associated with neurocognitive impairment in people living with HIV"

### Figure S1

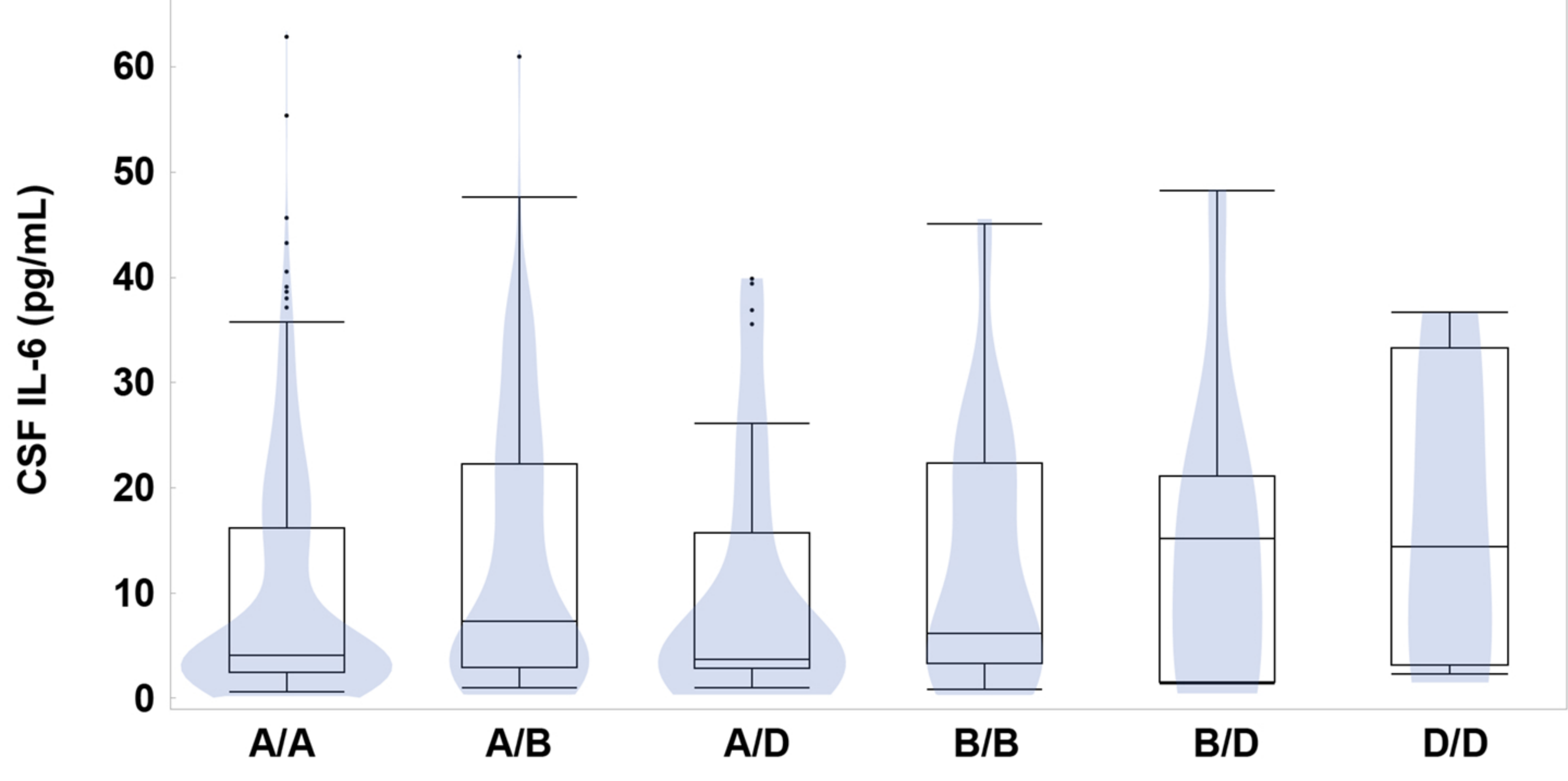
