## Supplementary material for "Genetic variations in *EIF2AK3* are associated with neurocognitive impairment in people living with HIV": Table S1

**Table S1.** Minor allele frequency of noncoding *EIF2AK3* SNVs

| | Number of minor alleles | | |
|---|---|---|---|
| | **0** | **1** | **2** |
| **rs6739095, T** | 58.3% | 35.1% | 6.6% |
| **rs1913671, C** | 58.3% | 34.8% | 6.6% |
| **rs11684404, C** | 60.6% | 33.0% | 6.4% |

SNV, single nucleotide variant
