## Supplementary material for "Genetic variations in *EIF2AK3* are associated with neurocognitive impairment in people living with HIV": Table S2

**Table S2.** Concordance of noncoding *EIF2AK3* SNVs

| | **rs6739095** | **rs1913671** | **rs11684404** |
|---|---|---|---|
| **rs6739095** | - | | |
| **rs1913671** | 99.6% | - | |
| **rs11684404** | 96.8% | 96.9% | - |

SNV, single nucleotide variant
