## Supplementary material for "Genetic variations in *EIF2AK3* are associated with neurocognitive impairment in people living with HIV": Table S3

**Table S3.** Characteristics of the targeted sequencing sub-cohort of participants included in targeted sequencing

| | All (n = 992) | Unimpaired (n = 636) | Impaired (n = 336) | *p* |
|---|---|---|---|---|
| Age, y (mean) | 43.7 | 44.0 | 43.6 | 0.27 |
| Sex, female | 22.6% | 22.7% | 22.4% | 0.93 |
| Ancestry | | | | **<0.0001** |
| European | 42.2% | 39.5% | 47.5% | |
| Admixed Hispanic | 9.8% | 6.6% | 15.7% | 0.004 |
| African ancestry | 45.9% | 52.3% | 33.8% | **<0.001** |
| AIDS diagnosis | 61.9% | 60.3% | 64.7% | 0.18 |
| HCV seropositivity | 26.8% | 29% | 22.6% | **0.03** |
| Nadir $CD4^+$ T-count, median (/µL) | 168 (41–295) | 175 (40–312) | 162 (47–265) | 0.14 |
| Current $CD4^+$ T cells, median (/µL) | 439 (283–615) | 441 (284–607) | 439 (281–624) | 0.33 |
| ART – current use | 74.9% | 71.6% | 81% | **0.001** |
| HCV seropositivity | 26.8% | 29% | 22.6% | **0.03** |

AIDS, acquired immunodeficiency syndrome; ART, antiretroviral therapy; HCV, hepatitis C virus
