## Supplementary material for "Genetic variations in *EIF2AK3* are associated with neurocognitive impairment in people living with HIV": Table S4

**Table S4.** Minor allele frequency of coding *EIF2AK3* SNVs

| | **Number of minor alleles** | | |
|---|---|---|---|
| **Allele** | **0** | **1** | **2** |
| **rs13045, A** | 58.8% | 34.5% | 6.8% |
| **rs867529, G** | 69.1% | 26.1% | 4.8% |
| **rs1805165, G** | 69.1% | 26.1% | 4.8% |

SNV, single nucleotide variant
