## Supplementary material for "Genetic variations in *EIF2AK3* are associated with neurocognitive impairment in people living with HIV": Table S5

**Table S5.** Concordance of coding *EIF2AK3* SNVs

| | **rs13045** | **rs867529** | **rs1805165** |
|---|---|---|---|
| **rs13045** | - | | |
| **rs867529** | 88.3% | - | |
| **rs1805165** | 88.3% | 100% | - |

SNV, single nucleotide variant
