## Supplementary material for "Genetic variations in *EIF2AK3* are associated with neurocognitive impairment in people living with HIV": Table S6

**Table S6**. Distribution of *EIF2AK3* haplotypes in the targeted sequencing sub-cohort

| | **A/A** | **A/B** | **A/D** | **B/B** | **B/D** | **D/D** |
|---|---|---|---|---|---|---|
| No. participants (%) | 583 (58.7%) | 245 (24.7%) | 97 (9.8%) | 48 (4.8%) | 14 (1.4%) | 5 (0.5%) |
| Age, y (median) | 44 | 43 | 42 | 43 | 47 | 45 |
| Race | | | | | | |
| European | 33.2% | 64.5% | 38.1% | 75% | 92.9% | 20% |
| Hispanic | 6.9% | 14.7% | 8.3% | 20.8% | 0% | 60% |
| Black | 59.9% | 20.8% | 53.6% | 4.2% | 7.1% | 20% |
| Sex, female | 24.7% | 20% | 22.7% | 10.4% | 14.3% | 40% |
| AIDS diagnosis | 60.4% | 60% | 62.3% | 56.3% | 71.4% | 100% |
| Nadir $CD4^+$ count, median ($/mm^3$) | 164 | 189 | 165 | 200 | 76 | 171 |
| ART-experienced | 85.6% | 82.6% | 82.3% | 85.1% | 85.7% | 80% |
| HCV seropositivity | 28.4% | 22% | 24% | 17% | 35.7% | 40% |
| NCI, n (%) | 199 (34.1%) | 162 (66.1%) | 61 (62.9%) | 27 (56.3%) | 7 (50%) | 2 (40%) |

AIDS, acquired immunodeficiency syndrome; ART, antiretroviral therapy; HCV, hepatitis C virus; NCI, neurocognitive impairment; NP, neuropsychiatric
