## Supplementary material for "Genetic variations in *EIF2AK3* are associated with neurocognitive impairment in people living with HIV": Table S7

**Table S7.** Global distribution of minor alleles of coding *EIF2AK3* SNVs

| | **Overall cohort** | **European** | **Asian** | **Latin** | **Black** |
|---|---|---|---|---|---|
| rs13045 | 33% | 33% | 47%–51% | 28%–39% | 9%–15% |
| rs867529 | 26% | 26% | 39%–46% | 21%–31% | 1%–7% |
| rs1805165 | 27% | 27% | 41%–47% | 21%–41% | 1%–7% |

SNV, single nucleotide variant
