## Supplementary material for "Genetic variations in *EIF2AK3* are associated with neurocognitive impairment in people living with HIV": Table S8

**Table S8.** Comparison of domain-specific deficit scores among groups categorized according to haplotypes

| | **Overall cohort** | **A/A** | **A/B** | **A/D** | **B/B** | **B/D** | **D/D** |
|---|---|---|---|---|---|---|---|
| Verbal | 0.27 ± 0.02 | 0.24 ± 0.02 | 0.35 ± 0.04 | 0.2 ± 0.06 | 0.4 ± 0.08 | 0.21 ± 0.15 | 0.2 ± 0.25 |
| Executive | 0.6 ± 0.02 | 0.54 ± 0.03 | 0.66 ± 0.05 | 0.73 ± 0.08 | 0.99 ± 0.12 | 0.61 ± 0.22 | 0.8 ± 0.36 |
| Learning | 0.67 ± 0.03 | 0.67 ± 0.04 | 0.7 ± 0.06 | 0.67 ± 0.09 | 0.87 ± 0.13 | 1.07 ± 0.24 | 0.7 ± 0.4 |
| Recall | 0.52 ± 0.03 | 0.58 ± 0.04 | 0.55 ± 0.06 | 0.61 ± 0.9 | 0.5 ± 0.13 | 0.89 ± 0.23 | 1.1 ± 0.39 |
| Working memory | 0.48 ± 0.02 | 0.47 ± 0.03 | 0.38 ± 0.04 | 0.52 ± 0.07 | 0.6 ± 0.1 | 0.54 ± 0.19 | 0.7 ± 0.31 |
| Motor | 0.49 ± 0.03 | 0.38 ± 0.03 | 0.54 ± 0.05 | 0.41 ± 0.08 | 0.44 ± 0.12 | 0.69 ± 0.23 | 1 ± 0.37 |
